# Glymphatic system impairment contributes to the formation of brain edema after ischemic stroke

**DOI:** 10.1101/2023.05.20.23290281

**Authors:** Juan Zhu, Jiaying Mo, Kewei Liu, Quanfeng Chen, Zheqi Li, Yihua He, Yuan Chang, Chuman Lin, Mingjia Yu, Yikai Xu, Xiangliang Tan, Kaibin Huang, Suyue Pan

**Affiliations:** Department of Neurology, Nanfang Hospital, Southern Medical University, Guangzhou, Guangdong, 510515, China; Department of Medical Imaging Center, Nanfang Hospital, Southern Medical University, Guangzhou, Guangdong, 510515, China

## Abstract

The blood-brain barrier (BBB) damage has traditionally been considered to determine the occurrence and development of post-stroke brain edema, a devastating and life-threatening complication. However, no treatment strategy targeting BBB damage has been proven clinically effective in ameliorating brain edema. Here, we found that the extravasation of protein-rich fluids into the brain was not temporally correlated with edema formation after middle cerebral artery occlusion (MCAO) in mice, as brain edema reabsorption preceded BBB closure. Strikingly, the time course of edema progression matched well with the glymphatic system (GS) dysfunction after MCAO. Pharmacological enhancement of the GS function significantly alleviated brain edema developed on day 2 after MCAO, accompanied by less deposition of Aβ and better cognitive function. Conversely, functional suppression of the GS delayed the absorption of brain edema on day 7 after MCAO. Moreover, patients with ischemic stroke revealed a consistent trend of GS dysfunction after reperfusion as MCAO mice, which was correlated with the severity of brain edema and functional outcomes. Collectively, these findings indicate that the GS is a key contributor to the formation of brain edema after ischemic stroke, and targeting the GS may be a promising strategy for the treatment of brain edema in ischemic stroke.

**One Sentence Summary:** The function of the glymphatic system is a key factor in determining the formation or resolution of brain edema after ischemic stroke

**Graphical Abstract:** 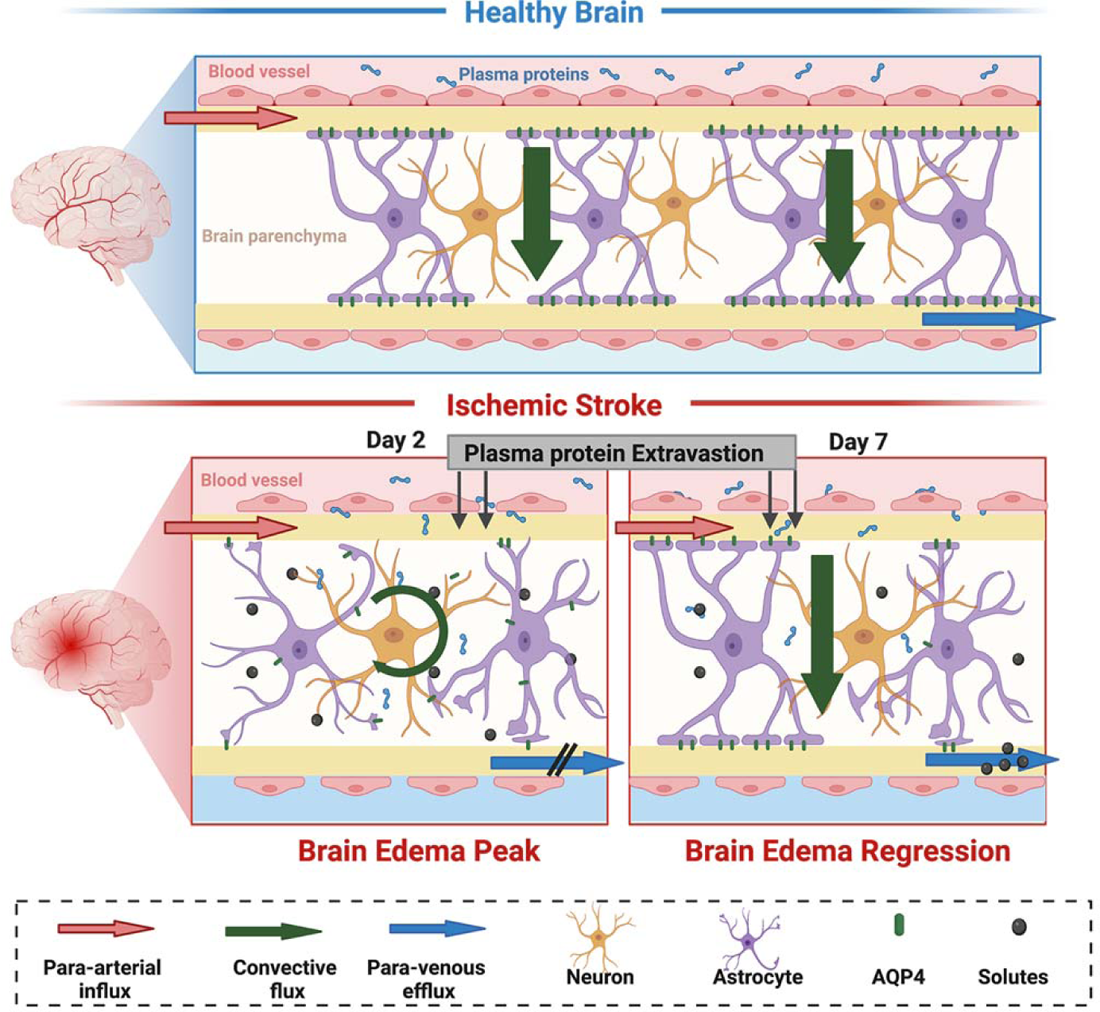

## INTRODUCTION

Stroke is the second highest cause of death worldwide and the main cause of long-term disability(*1*), with ischemic stroke accounting for the majority of cases(*2*). Brain edema is characterized by fluid accumulation within the brain tissue, leads to secondary neurological deterioration and irreversible neuron damage, and is the main culprit responsible for the high mortality and disability in stroke patients(*3*). However, the treatment options for brain edema remain limited and suboptimal. Therefore, it is imperative to explore the mechanisms responsible for brain edema formation in ischemic stroke for targeted therapy.

Generally, brain edema is divided into two distinct stages: the early stage of cytotoxic and ionic edema with an intact blood-brain barrier (BBB), and the later stage of vasogenic edema with impaired BBB(*4*). Previous studies mostly focus on the role of BBB damage in the occurrence and progression of brain edema. Several pathophysiologic processes, including the disruption of tight junctions (TJs)(*5*), inflammatory responses(*6*), and the loss of homeostatic ionic gradients(*4*), are thought to contribute to brain edema after stroke by increasing BBB permeability. In previous studies(*7–9*), including ours(*10*), BBB leakage after ischemic stroke was found to be continuous and biphasic, with the first phase of enhanced permeability occurring 4-6 hours after reperfusion and the second phase occurring 7 days after reperfusion. However, brain edema induced by ischemic stroke peaked on day 2 and resolved on day 7 in animal models(*11–13*). These data raise an apparent paradox that, in the late stage of ischemic stroke (i.e., on day 7 after reperfusion), the brain edema begins to subside, while the BBB remains to be leaky or opening(*14*). Currently, it is unclear how the excess fluid in brain edema is cleared, given that the brain lacks a conventional lymphatic system.

The glymphatic system (GS) is a newly discovered solute and waste clearance system from the brain parenchyma through promoting the exchange of cerebrospinal fluid (CSF) and interstitial fluid (ISF), which consists of a brain-wide paravascular space (PVS)(*15*). Anatomically, GS is made up of three key parts: para-arterial influx of CSF into the brain parenchyma, convective fluid flow facilitated by high-polarized aquaporin-4 (AQP4) water channels, and para-venous or -nerve efflux of ISF into the meninges and deep cervical lymph nodes (dCLNs)(*16, 17*). It can be inferred that when the GS transports CSF inflow into cerebral parenchyma is enhanced and/or the outflow of ISF is blocked, abnormal accumulation of cerebral tissue fluid may occur and lead to brain edema(*14*). A recent study described that the influx of CSF into the brain tissue drove acute tissue swelling within minutes of ischemic stroke, suggesting that ionic edema is caused by the influx of CSF rather than intravascular fluid traditionally assumed(*18*). However, the role of the GS in the later stage of brain edema when the BBB is opening has not been addressed. In particular, if the GS is involved, does it aggravate brain edema by promoting CSF inflow, or alleviate edema by mediating ISF outflow?

In this study, we investigated the contribution of GS to the formation of brain edema after ischemic stroke. We found that the BBB leakage was not synchronized with brain edema evolution after transient middle cerebral artery occlusion (MCAO) in mice, as BBB leakage was detected not only during the expanding (peak) period of edema (day 2) but also during the absorption phase of edema (day 7). Strikingly, the time course of brain edema progression matched well the GS dysfunction after MCAO. Pharmacological enhancement of the GS function significantly alleviated brain edema developed on day 2 after MCAO, accompanied by less deposition of Aβ and better cognitive function. Conversely, functional suppression of the GS delayed the absorption of brain edema on day 7 after MCAO and led to more deposition of Aβ and poorer cognitive function. Moreover, patients with ischemic stroke revealed a consistent trend of GS dysfunction after reperfusion as MCAO mice, which was correlated with the severity of brain edema and functional outcomes. These findings unveil a previously unrecognized role of GS in the formation of brain edema after ischemic stroke, and provide GS as a novel therapeutic target for brain edema.

## RESULTS

### 1. The evolution of brain edema is inconsistent with BBB leakage after transient focal brain ischemia

Cranial MRI was utilized to observe the dynamic evolution of brain edema within the first week after transient focal brain ischemia. MCAO-induced swelling of brain tissue was examined by HEV% and MLS. As shown, the values of HEV% and MLS distinctly increased during the acute phase, and peaked on day 2 after reperfusion, then gradually re-normalized, reaching the minimum on day 7 (**Fig. 1A-C**). This temporal profile of brain swelling indicated that vasogenic edema induced by MCAO peaked on day 2 and almost resolved on day 7, which was in agreement with the previous reports(*11–13*). Moreover, the rADC value decreased during the acute phase and reached the minimum on day 2 after reperfusion, then slowly recovered on day 7 (**Fig. 1D**), indicating that there was a coexistence of cytotoxic edema and vasogenic edema during the acute phase. Electronic microscopic changes in cerebral cortical capillaries after MCAO revealed that astrocyte edema was prominent on day 1 and day 2, and subsided on day 7 (**Fig. 1E**), which was accompanied by a significant widening of the basement membrane. Collectively, these results suggested that brain edema peaked on day 2 after MCAO, and subsided on day 7.

**Fig. 1.**
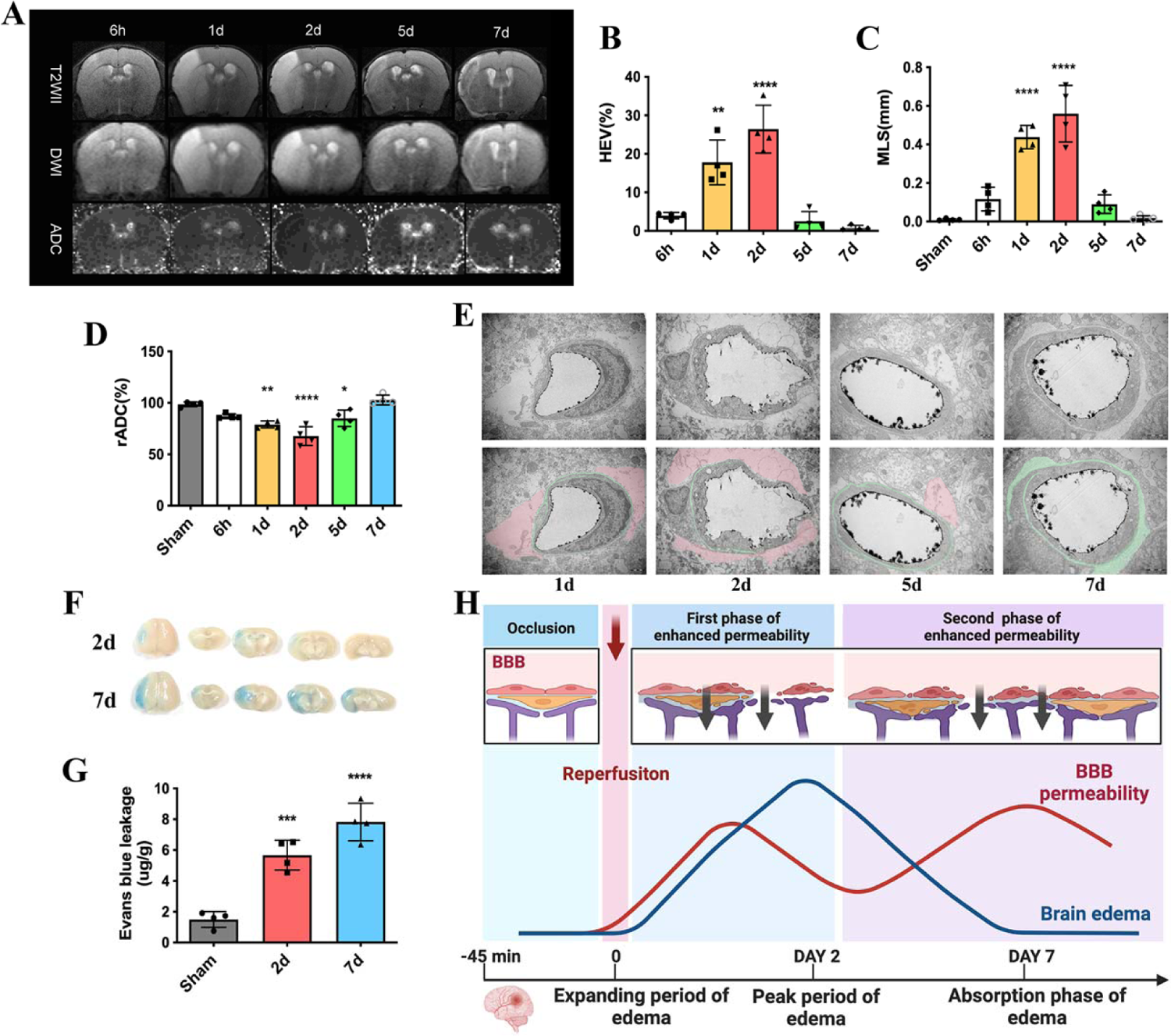
Short title of the first figure. Brain edema subsides when BBB leakage increased on day 7 after transient focal brain ischemia. (A) Representative MRI images of T2 and ADC for edema analysis at 6h, 1d, 2d, 5d and 7d post reperfusion. (B-D) Quantitative analysis of HEV%, MLS and ADC at corresponding time points showed that brain edema peaked on day 2 after MCAO, and subsided on day 7. (E) Representative EM images of individual capillary at different time points after reperfusion. Red indicates the swelling degree of astrocytes. The basement membrane is marked in green. (F-G) Representative images and quantitative analysis of EB extravasation showed that BBB leakage increased 7 d after MCAO with edema subsided. (H) The diagram shows that in the late stage of ischemic stroke (i.e., on day 7 after reperfusion), the brain edema begins to subside, while the BBB remains to be leaky or opening. *p < 0.05 vs. sham group.

Our previous research found that the BBB presented a continuous and biphasic opening for the first week after MCAO, with the first permeability peak on day 2 and a stronger second peak on day 7(*10*). Apparently, under our experimental conditions, the BBB leakage was not synchronized with brain edema progression, as BBB leakage was detected not only during the expanding (peak) period of edema (day 2), but also during the absorption phase of edema (day 7) after MCAO (**Fig. 1F-G**). This intriguing result raised an important question: why did brain edema subside when the BBB leakage was still increasing on day 7 after MCAO (**Fig. 1H**)?

### 2. Paravascular CSF influx is severely damaged on day 2 after MCAO and restored on day 7

To answer the question above, we first evaluated the effect of MCAO on the GS function, focusing on the peak period of brain edema (day 2 after MCAO) and the regression period (day 7 after MCAO). CSF fluorescence tracer EB dye (MW 960 Da) was slowly injected into the subarachnoid CSF of the cisterna magna (**Fig. 2A**), and 30 min post injection, EB penetration into the brain parenchyma was evaluated ex vivo by fluorescence microscopy(*15*). Compared with sham controls, EB dye was broadly dispersed within the ischemic hemisphere on day 2 and day 7, and specifically, the tracer distribution was consistent with the infarcted lesion, suggesting that this distribution probably resulted from the disrupted BBB, which made it difficult to observe the ipsilateral cortical tracer influx. We instead observed that CSF tracer penetration in the contralateral cortex was dramatically decreased along the PVS and across the pia mater surface on day 2, compared with day 7 (**Fig. 2B-C**).

**Fig. 2.**
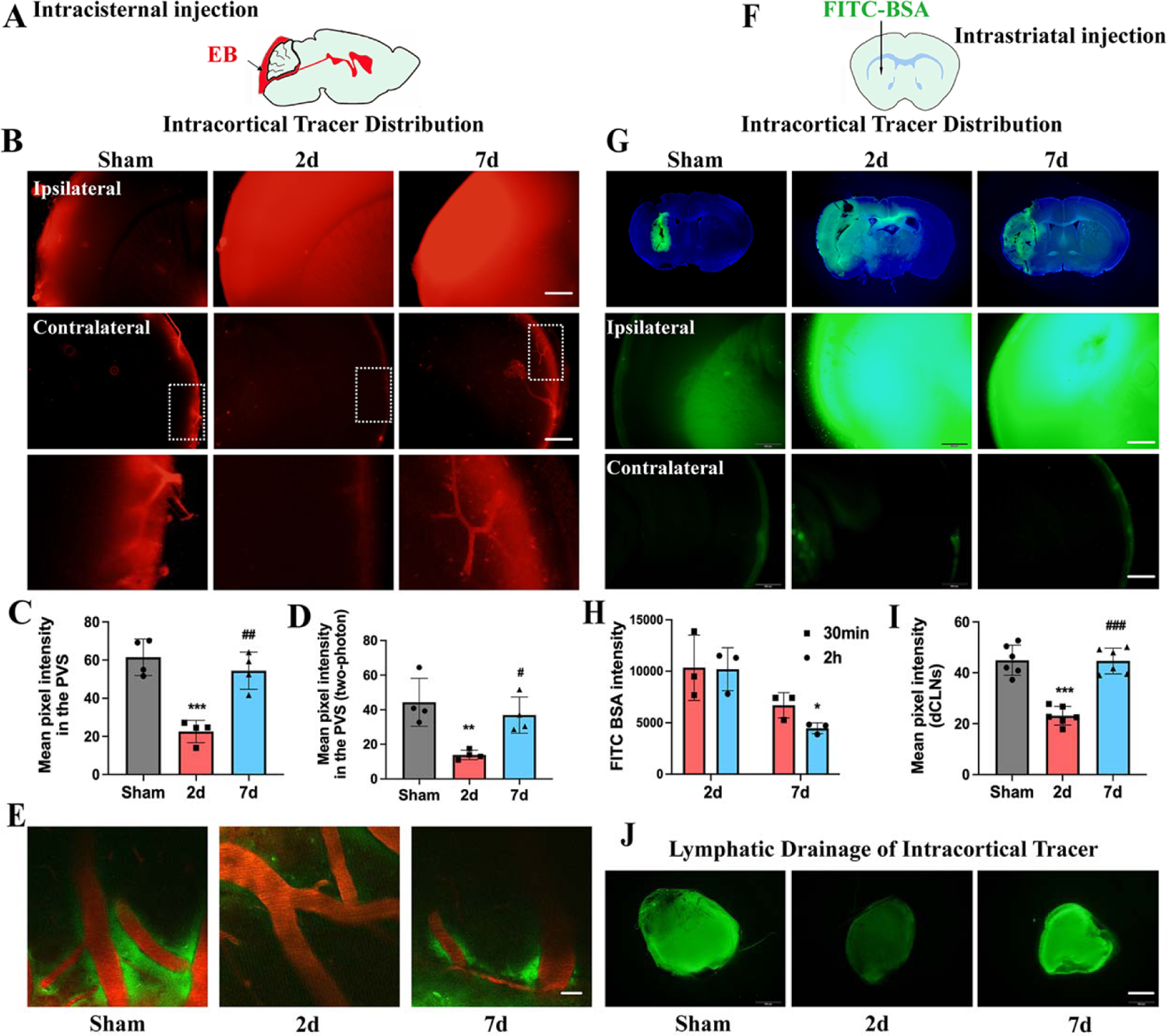
Glymphatic system function is severely impaired on day 2 after MCAO and restored on day 7. **(A)** Schematic depiction of intracisternal administration of CSF tracer. **(B)** Representative images (2mm thick, observation location: bregma −1 mm) of fluorescence tracer (red, EB) in coronal brain sections (upper panel, scale bar: 500 μm) and higher-magnification images for the PVS (lower panel, derived from boxes in upper panel). **(C)** Quantitative analysis of the mean pixel intensity of the tracer in the PVS. **(D-E)** Representative two-photon fluorescence images of the cerebral vasculature and the distribution of the tracer (observation location: 100 μm below brain surface). Red, cerebral vasculature; green, cerebrospinal fluid tracer. (Scale bar: 30 μm) **(F)** Schematic depiction of intrastriatal administration of CSF tracer. **(G)** Representative images (bregma −1 mm) of fluorescence tracer (FITC-BSA) injected into the brain parenchyma (Scale bar: 500 μm). **(H)** Quantitative analysis of the mean pixel intensity of the tracer in the brain parenchyma. **(I-J)** Representative images and quantitative analysis of the fluorescence intensity of FITC-BSA drainage into the dCLNs (Scale bar: 500 μm). *p < 0.05 vs. sham group. ^#^p < 0.05 vs. 2d group.

Due to perfusion fixation, ex vivo post-mortem imaging may affect the tracer uptake patterns and the data interpretation(*19*). Hence, we performed in vivo 2-photon microscopy to evaluate the function of GS influx following intracisternal injection of FITC-dextran (MW 70 kDa; **Fig. 2D-E**). The cerebral vasculature was visualized through a thinned cranial window after tail-vein injection of RITC-dextran (MW 40 kDa). Thirty minutes after intracisternal injection, para-arterial CSF tracer influx was observed at the cortical surface of sham-operated mice. However, only a weak CSF tracer fluorescent signal appeared in the PVS of mice on day 2 after MCAO. On day 7 after MCAO, CSF tracer penetration was markedly increased compared to that on day 2. These results indicated that paravascular CSF influx was severely impaired on day 2 after MCAO, and restored on day 7.

### 3. Glymphatic clearance is significantly impaired on day 2 and recovered on day 7 after MCAO

Glymphatic influx only partly reflects the function of the GS, therefore we next measured the glymphatic clearance after MCAO. FITC-BSA was injected into the striatum and the clearance of the tracers was measured 30 min and 2 h later (**Fig. 2F**). At 2 h after intrastriatal injection of FITC-BSA, sham mice showed restricted fluorescent intensity within the brain parenchyma, with strong fluorescent intensity within the dCLNs (**Fig. 2G-J**). In contrast, mice on day 2 after MCAO displayed a higher percentage of fluorescent signal within the striatum and a lower percentage of fluorescent signal within the dCLNs, indicating that the clearance of solutes from the brain was impaired on day 2 after MCAO. This phenomenon almost recovered on day 7 after MCAO, suggesting that the glymphatic clearance was more efficient on day 7 than that on day 2 after MCAO.

To further unambiguously demonstrate the tracer uptake patterns, we used in vivo 2-photon microscopy to detect the glymphatic clearance of RITC-dextran (MW 40 kDa) after MCAO. As shown in **Fig. 3**, there was no significant difference in RITC-dextran intensity at 5 min post-injection between the two groups of day 2 and day 7 after MCAO, and the RITC-dextran signal intensity of the two groups kept decreasing from 5 to 60 min post-injection. However, from 15 min to 60 min following RITC-dextran injection, the RITC-dextran signal intensity on day 2 after MCAO was significantly higher than that on day 7, indicating that the glymphatic clearance on day 2 after MCAO was significantly weaker than that on day 7.

**Fig. 3.**
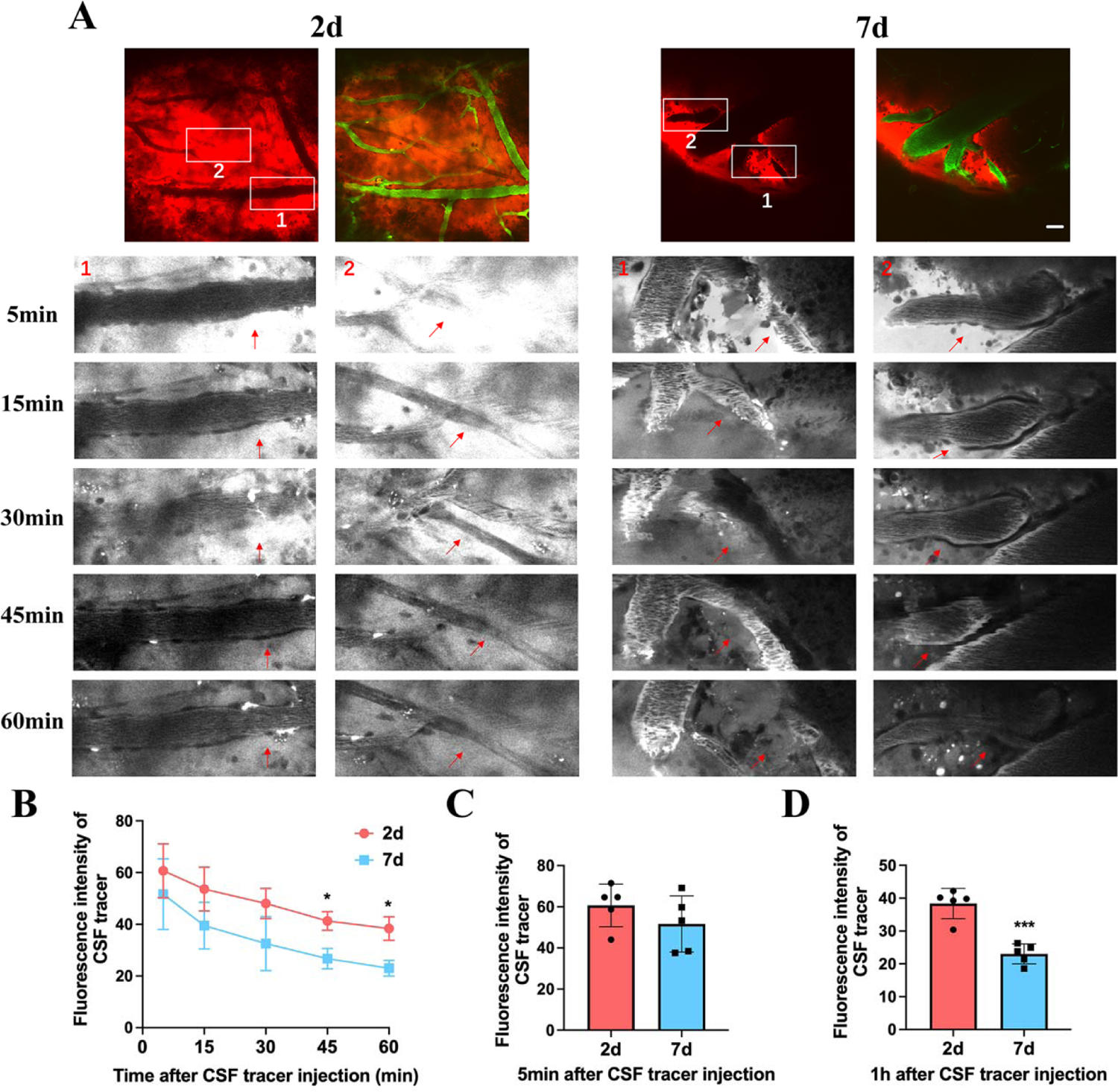
The glymphatic clearance on day 2 is severely impaired compared with day 7, as demonstrated by in vivo two-photon imaging. **(A)** Representative two-photon fluorescence images of the cerebral vasculature and the distribution of the tracer over time. To visualize GS clearance in the mouse cortex, a 40 kDa dextran conjugated to Rhodamine B was injected into the brain parenchyma at about 200 μm depth via a micropipette. A 70 kDa dextran conjugated to FITC was injected i.v for visualization of the brain vasculature (Scale bar: 40 μm). **(B-D)** Quantitative analysis of fluorescence intensity of Rhodamine B conjugated dextran, showing a dramatic impairment in the dextran clearance along the vessel. *p < 0.05 vs. 2d group.

### 4. Glymphatic transport of Gd-DTPA on day 2 after MCAO in comparison to day 7

The limitations of the above fluorescence-based imaging modalities prevented the direct assessment of brain-wide CSF-ISF flow dynamics. We next used contrast-enhanced MRI to visualize the brain-wide subarachnoid CSF-ISF exchange and ISF efflux after MCAO. Dynamic T1WIs were acquired continuously for 6 h after injecting a low molecular weight compound Gd-DTPA (6 μL, 1.6 μL/min, MW 938 Da) into the cisterna magna. As reported in previous MRI studies(*20, 21*), the serial T1WIs over 6 h clearly exhibited the anatomical routes of glymphatic influx from the cisterna magna and clearance from brain parenchyma (**Fig. 4**), including 1) within 30 min, the Gd-DTPA traveled along the basilar artery, passed through the pituitary and along the olfactory arteries, and arrived at the olfactory bulb; 2) contrast enhancement was observed in the whole brain at 2 h after Gd-DTPA injection and displayed partial difference at 4 h; 3) contrast enhancement was decreased at 6 h after Gd-DTPA injection.

**Fig. 4.**
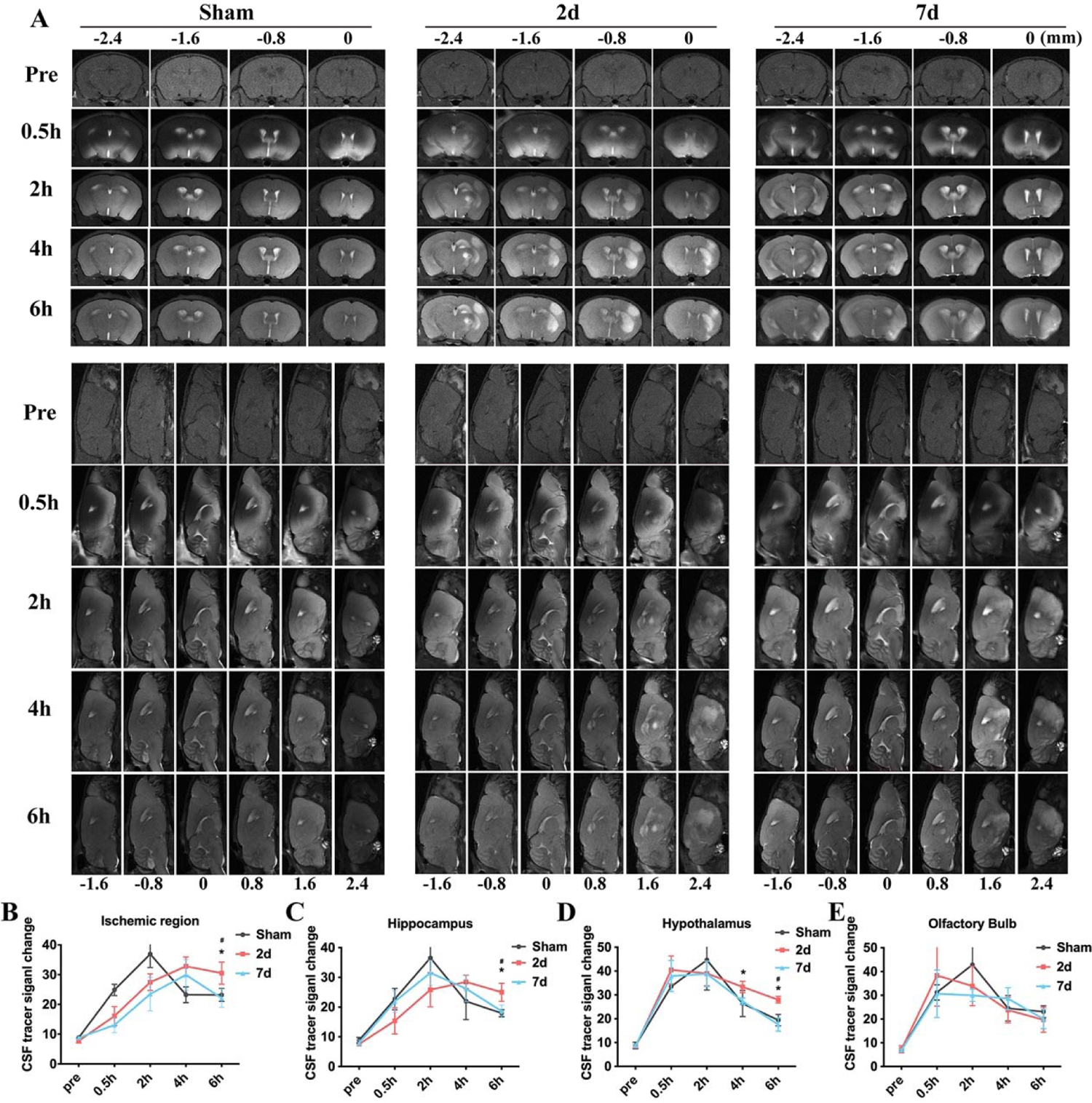
Glymphatic system function on day 2 is severely impaired compared with day 7, as demonstrated by contrast-enhanced MRI. **(A)** Representative MRI coronal (top) and sagittal (bottom) images of early influx and late efflux of the contrast agent Gd-DTPA (MW 938 Da) along the paravascular pathway over 6 hours (i.e., pre, 0.5, 2, 4, 6 h). **(B-E)** Quantitative analysis of T1WI changes of the responding regions. *p < 0.05 2d vs. 7d group, ^#^p < 0.05 2d vs. sham group.

Of note, although the evolution of Gd-DTPA signal intensity at the peak of brain edema (day 2 after MCAO) exhibited similar time courses with the regression of brain edema (day 7 after MCAO) within the first 4 h in the brain regions examined (e.g., ischemic region and hypothalamus), the Gd-DTPA appeared to enter the hippocampus more rapidly when brain edema subsided (day 7 after MCAO), exhibiting a peak signal at 2 h in advance. Moreover, the mice on day 2 after MCAO exhibited higher residual intensity compared with the mice on day 7 at these brain regions over the 6-h infusion. Consistent with previous fluorescent imaging data, the overall changes of Gd-DTPA on day 2 after MCAO versus day 7 reflected a slower clearance rate during CSF-ISF exchange.

### 5. AQP4 polarization is significantly impaired on day 2 after MCAO and partly recovered on day 7

The function of GS is highly dependent upon the localization of the water channel AQP4 to the endfeet of astrocytes that surround brain microvasculature, which is defined as a polarized distribution of AQP4(*22*). Therefore, we detected the changes of AQP4 localization to the cerebral microvasculature on brain slices collected from MCAO mice. Using immunohistochemistry, we found that the polarization of AQP4 around the microvascular structures was significantly decreased on day 2 after MCAO across the infarcted core and peri-infarct regions (**Fig. 5**). However, the AQP4 polarization in these regions was significantly higher on day 7 after MCAO than on day 2, supporting the results that the GS function was damaged on day 2 after MCAO and partly restored on day 7. Overall, the above results demonstrated a close temporal correlation between the evolution of brain edema after MCAO and the functional status of GS.

**Fig. 5.**
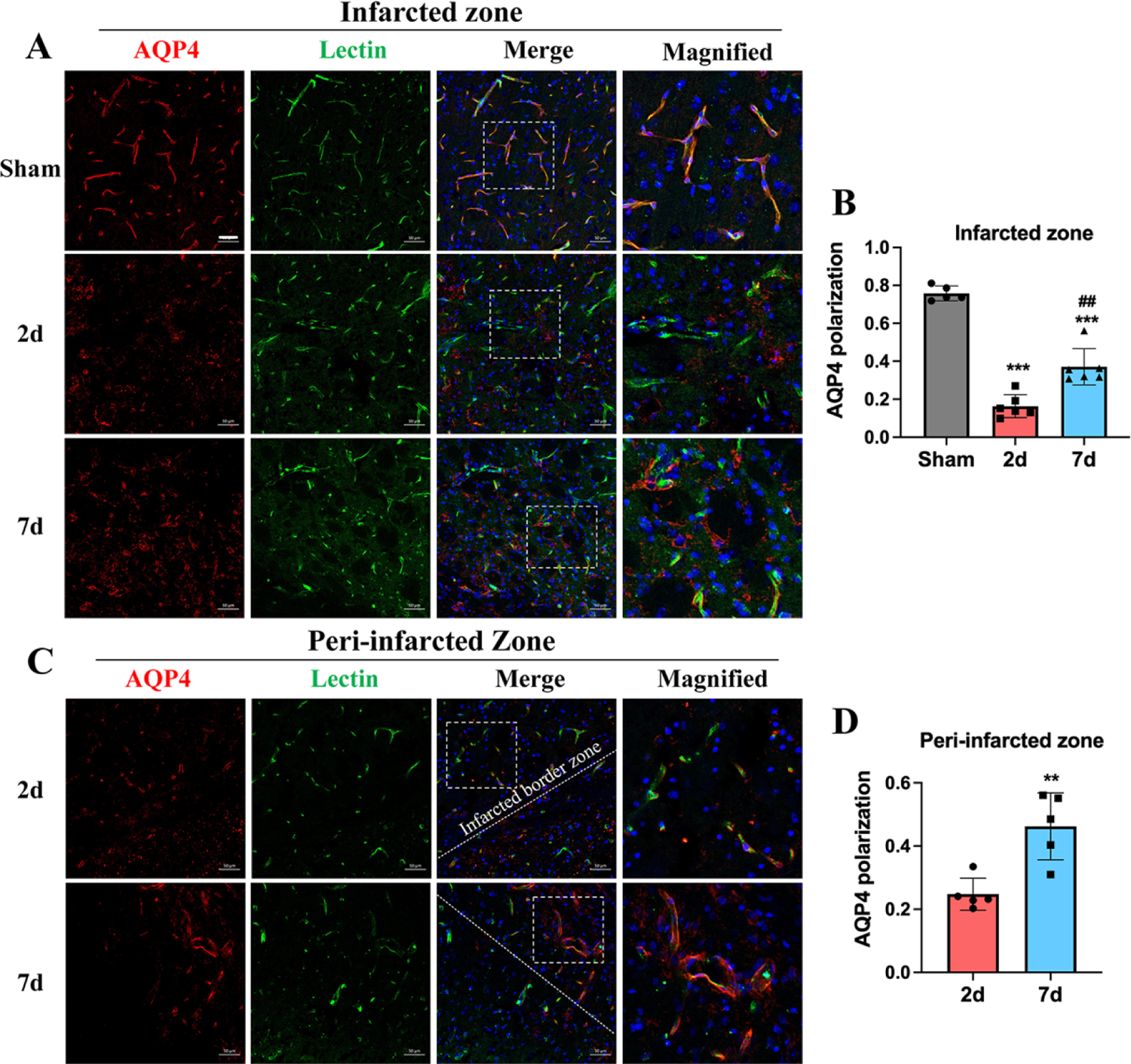
Analyses of AQP4 polarization at the infarcted zone and peri-infarcted zone after MCAO. **(A)** Representative confocal images perivascular polarization of AQP4 at infarcted zone on 2 d and 7 d after MCAO. Red is AQP4, green is vascular staining. **(B)** Quantitative analysis of perivascular polarization of AQP4 showed that AQP4 polarization on day 2 was severely impaired compared with day 7. **(C-D)** Representative confocal images and quantitative analysis of AQP4 polarization in the peri-infarcted zone (Scale bar: 50 μm). *p < 0.05 vs. 2d group.

### 6. Enhancement of GS function alleviates brain edema on day 2 after ischemic stroke

In the above experiments, we have shown that brain edema subsided with the recovery of GS function on day 7 after MCAO when the BBB permeability kept increasing. We speculated that the recovery of GS function on day 7 after MCAO contributed to the clearance of excessive intravascular fluids resulting from the leaky BBB, thus leading to the regression of brain edema. To test this hypothesis, we suppressed or enhanced GS function and examined the corresponding change in brain edema after MCAO. Xie et al.(*23*) found that adrenergic inhibition by injecting a cocktail of adrenergic receptor antagonists promoted GS transporting function in mice.

Therefore, we assessed the effect of adrenergic inhibition on post-MCAO GS transport using ex vivo imaging, and found that CSF tracer influx was significantly increased after adrenergic inhibition on day 2 after MCAO (**Fig. 6A-B**). Besides, the fluorescence intensity in the dCLNs of mice treated with adrenergic receptor antagonists was significantly stronger than that in the control mice (**Fig. 6C-E**). These results supported that adrenergic inhibition indeed improved the GS transporting function.

**Fig. 6.**
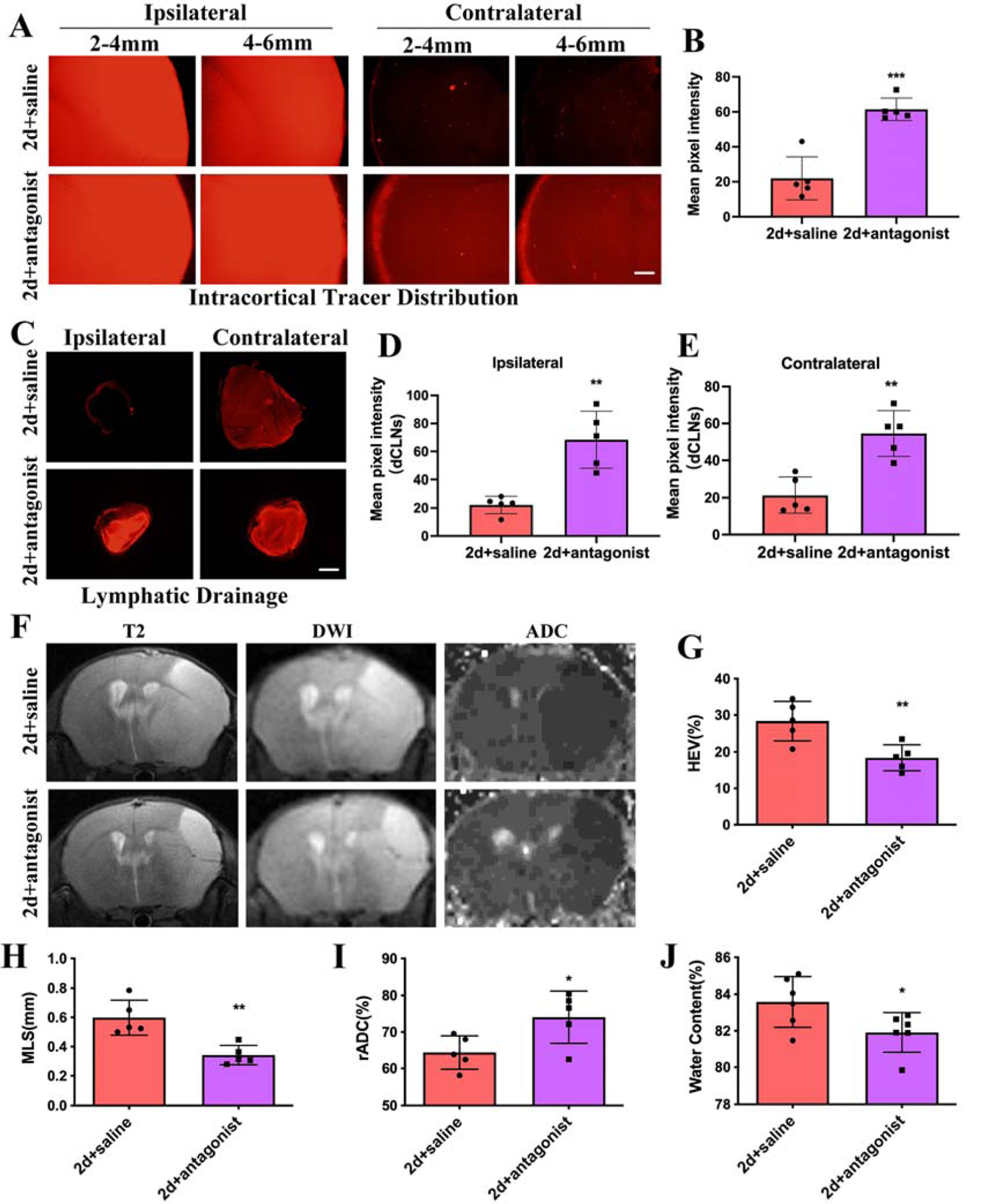
Brain edema significantly alleviates on 2d after MCAO by promoting GS function using a cocktail of adrenergic receptor antagonists. **(A)** Representative images of fluorescence tracer (red, EB) in coronal brain sections (Scale bar: 500 μm). **(B)** Quantitative analysis of the mean pixel intensity of the tracer in the PVS. **(C-E)** Representative images and quantitative analysis of the fluorescence intensity of tracer drainage into the dCLNs (Scale bar: 500 μm). **(F)** Representative MRI images of T2 and ADC for edema analysis on 2d post reperfusion. **(G-I)** Quantitative analysis of HEV%, MLS and ADC showed that brain edema significantly alleviated after GS function promotion. **(J)** Water content decreased significantly after GS function promotion. *p < 0.05 vs. 2d+saline group.

We then examined the impact of adrenergic inhibition on brain edema. Adrenergic inhibition significantly reduced the values of HEV% and MLS, and increased the rADC values on day 2 after MCAO (**Fig. 6F-I**). Moreover, compared with the vehicle group, the water content significantly decreased in the group of adrenergic inhibition (**Fig. 6J**). These results suggested that promoting the GS function significantly alleviated brain edema on day 2 after MCAO.

### 7. Suppression of GS function augments brain edema on day 7 after ischemic stroke

We then assessed the effect of adrenergic stimulation on post-MCAO GS transport during the regression period of brain edema. As shown, CSF tracer influx was significantly decreased after adrenergic stimulation on day 7 after MCAO (**Fig. 7A-B**), which reflected a decrease of tracers at the edge of brain slices and the PVS surrounding microvessels. Moreover, the fluorescence intensity in the dCLNs in the adrenergic stimulation group was significantly weaker than that in the control group (**Fig. 7C-E**). These results suggested that adrenergic stimulation inhibited GS transporting function.

**Fig. 7.**
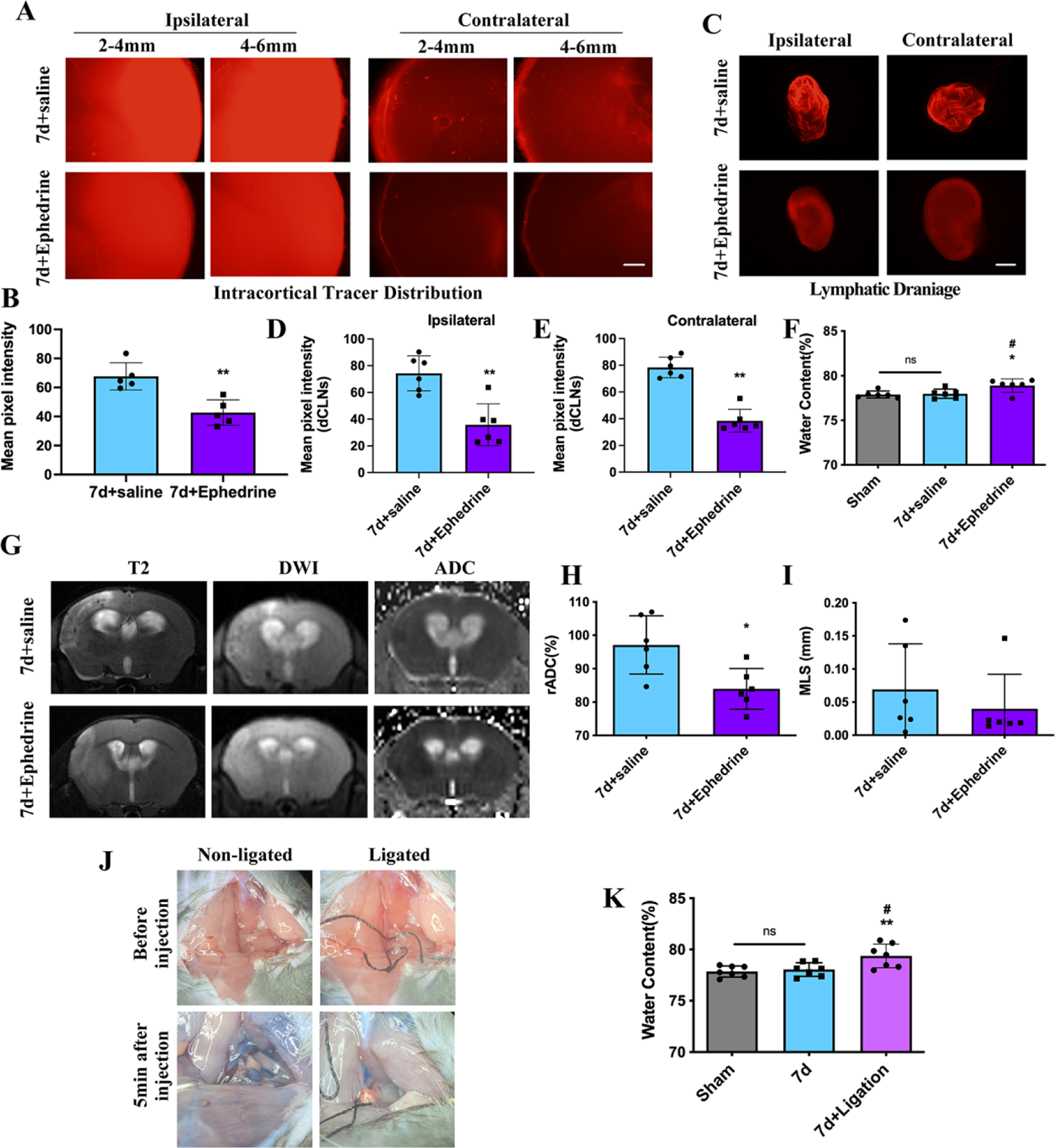
Functional suppression of the GS delays the absorption of brain edema on day 7 after MCAO. **(A)** Representative images of fluorescence tracer (red, EB) in coronal brain sections (Scale bar: 500 μm). **(B)** Quantitative analysis of the mean pixel intensity of the tracer in the PVS. **(C-E)** Representative images and quantitative analysis of the fluorescence intensity of tracer drainage into the dCLNs (Scale bar: 500 μm). **(F)** Quantitative analysis of water content showed that suppression of the GS significantly increased water content of brain tissue on day 7 after MCAO. **(G-I)** Representative MRI images of T2 and ADC for edema analysis on 7d post reperfusion, quantitative analysis of ADC showed that brain edema occurred after GS function suppression. **(J-K)** Water content increased in mice with dCLNs ligation 7 days after MCAO. *p < 0.05 vs. 7d+saline group.

As expected, adrenergic stimulation significantly increased water contents of brain tissue on day 7 after MCAO (**Fig. 7F**) and reduced the values of rADC on day 7 after MCAO, compared with the vehicle group, while the change of MLS values was not statistically significant (**Fig. 7G-I**). On this basis, we used ligation of the dCLNs, another method to inhibit the GS function, and found that the water contents were also significantly increased in mice with dCLNs ligation 7 d after MCAO (**Fig. 7J-K**). Collectively, these results indicated that the progression and resolution of brain edema coincided with the impairment and recovery of GS function, and that intervention of the GS function could significantly modulate the extent of brain edema, suggesting that GS transport may play a decisive role in the progression of brain edema after ischemic stroke, even when BBB leakage exists.

### 8. Improving GS function significantly mitigates cognitive impairment and deposition of Aβ after MCAO

The GS is not only an important pathway for fluid exchange and clearance in the brain but also crucial for metabolic waste clearance(*24*). Impairment of glymphatic clearance leads to the accumulation of protein aggregates and cognitive decline in traumatic brain injury(*25*) and neurodegenerative diseases(*26*). We thus evaluated the effect of GS function on cognitive function after MCAO. As shown in **Fig. 8**, in the Morris water maze test, the times of crossing the platform and the time in the target quadrant increased in mice treated with adrenergic receptor antagonists, which was accompanied by significantly reduced deposition of Aβ. Inversely, mice treated with adrenergic receptor agonists ephedrine showed fewer times crossing the platform, less time of staying in the target quadrant, and increased deposition of Aβ, compared with the vehicle-treated group. These data suggested that alleviating GS dysfunction by adrenergic inhibition may facilitate the removal of metabolic wastes from the brain and thus improve cognitive function after MCAO.

**Fig. 8.**
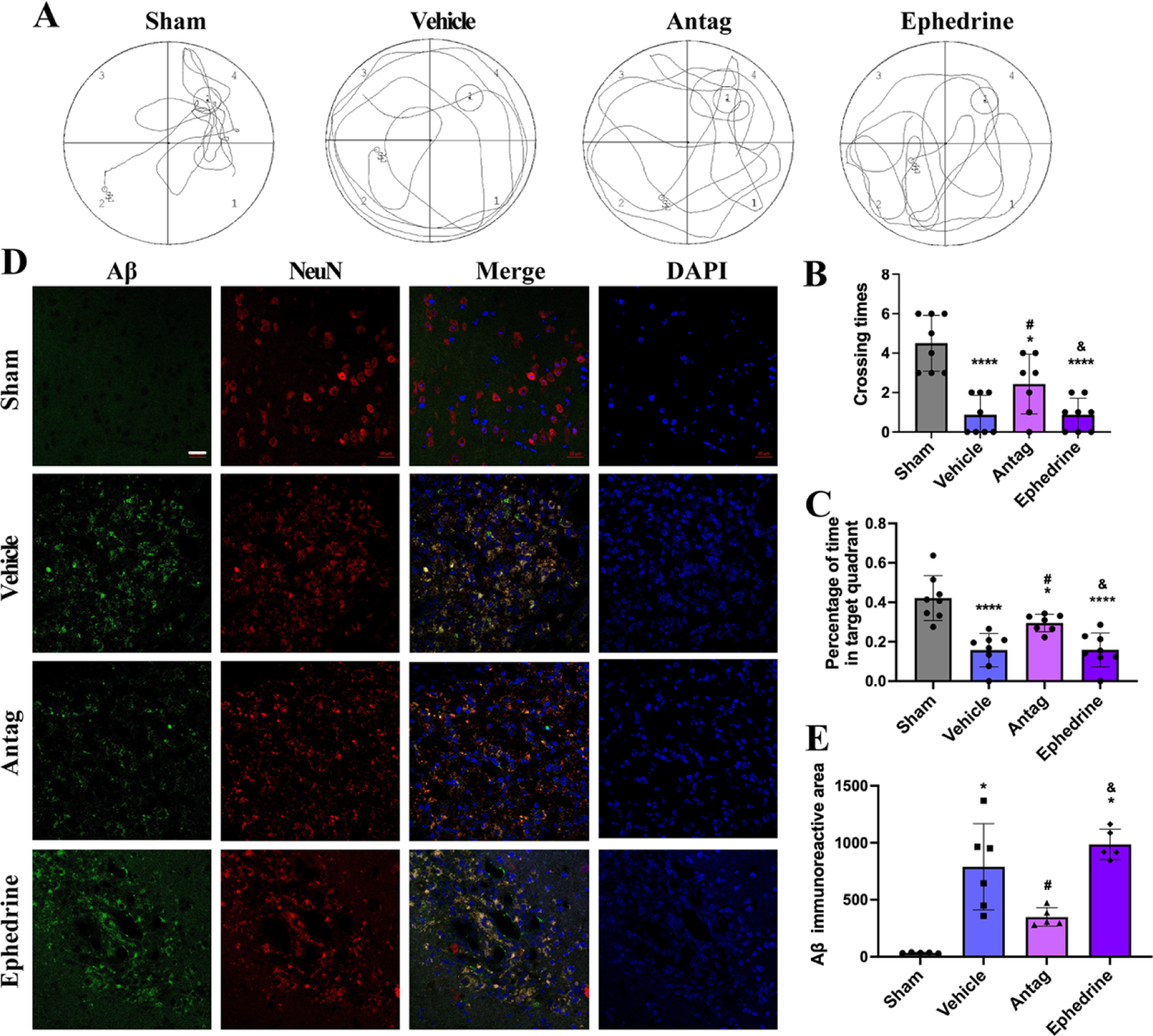
GS function promotion significantly mitigates cognitive impairment after MCAO. **(A-C)** The effect of GS function on post-stroke spatial memory and learning deficits assessed by Morris water maze analysis, including the frequency of crossing the platform area, and percentage of time spent in the target quadrant (Q4) during the probe trial. **(D-E)** Representative images and quantitative analysis of Aβ-deposited neurons in the infarcted hemisphere (red: NeuN; green: Aβ; blue: DAPI). Scale bars: 20 μm. *p < 0.05 vs. sham group; ^#^p < 0.05 vs. vehicle group; ^&^p < 0.05 vs. Antag group. Antag: antagonists.

### 9. Glymphatic clearance is significantly impaired from day 1 to day 3 and recovered on day 7 after acute ischemic stroke in human

To determine whether the pattern of glymphatic dysfunction seen in mice after stroke was clinically relevant, we measured the glymphatic clearance function using DTI-ALPS index(*27*) (**Fig. 9A-B**) in 18 patients with acute ischemic stroke and achieved recanalization after EVT (**Supplementary Table 1**). All the 18 patients had a DTI examination at 24 ± 4 hours (TIME 1) after EVT, 10 patients underwent a second DTI imaging at 72 ± 12 hours (TIME 2), and 5 patients underwent a third DTI imaging on day 7 ± 1 (TIME 3; **Supplementary Fig. 1**). In TIME 1 and TIME 2, the ALPS index of the infarcted side was significantly lower than that of the contralateral side, but the difference evanished in TIME 3 (**Fig. 9C**, **Supplementary Table 2**). A two-by-two comparison of the primary data showed that the ALPS index decreased from TIME1 to TIME2, and then increased from TIME 2 to TIME 3 for both the infarcted and contralateral sides (all *P* < 0.05). Besides, the overall difference in ALPS index was statistically significant among time (*P* = 0.017 for TIME 1 vs. TIME 2; *P* = 0.002 for all the three time points; **Supplementary Fig. 2, Table 3** and **Table 4**). Together, these data indicated that patients with acute ischemic stroke had significantly impaired glymphatic clearance function from day 1 to day 3 after reperfusion, especially on day 3, and almost recovered on day 7.

**Fig. 9.**
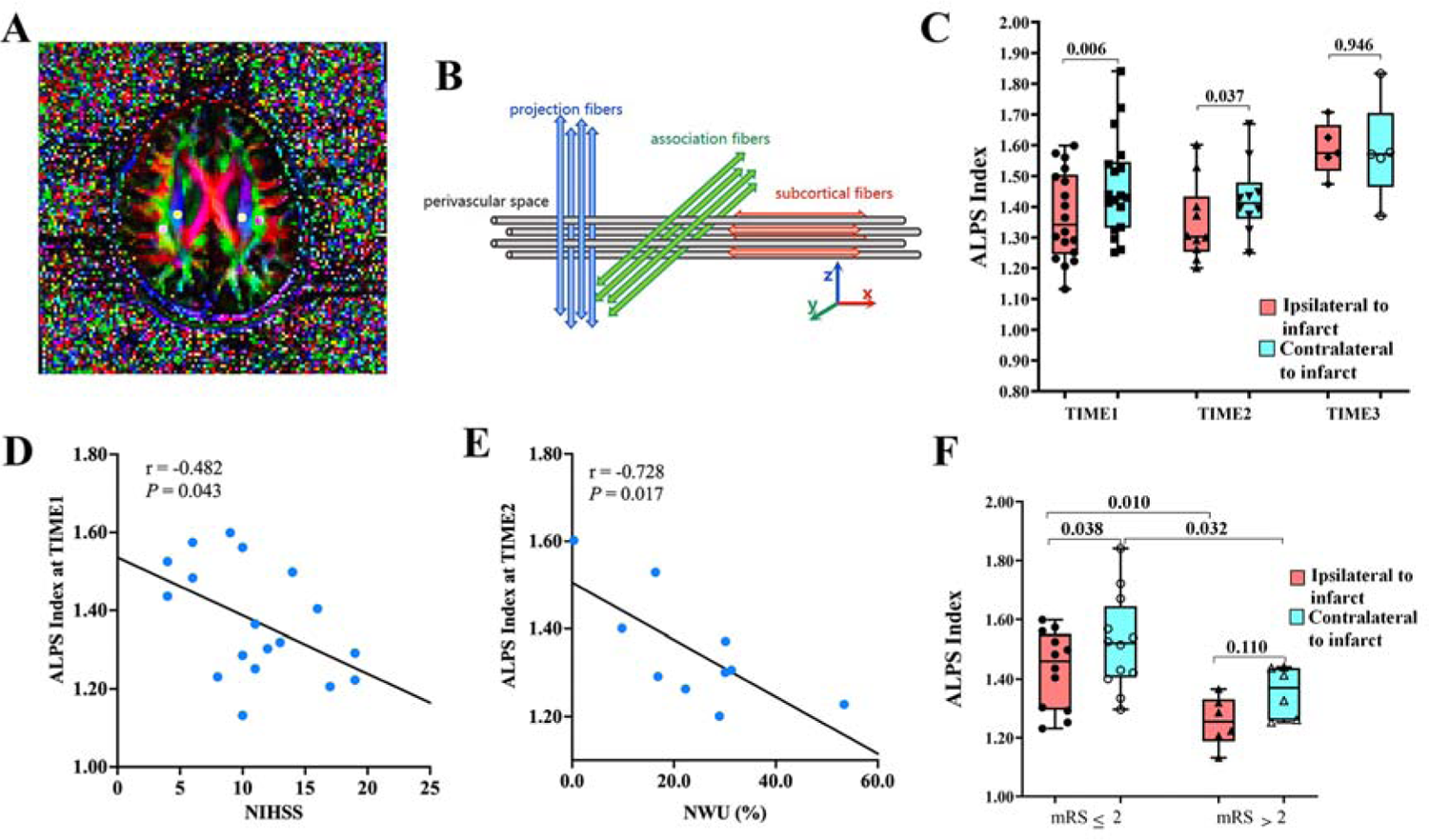
Glymphatic clearance is significantly impaired after acute ischemic stroke in humans and is associated with brain edema and functional outcomes. **(A)** Four 6-mm-diameter ROIs were placed in the area of bilateral projection and association fibers on color-code FA maps. **(B)** In the image coordinate, the perivascular space, projection fibers, and association fibers are nearly perpendicular to each other. **(C)** Boxplot showed interhemispheric differences in the ALPS index at different time points. **(D)** Scatterplots with regression lines showed the correlations of the ALPS index with NIHSS. **(E)** Scatterplots with regression lines showed the correlations of the ALPS index with NWU of the ischemic lesion. **(F)** Box plots showed the difference in infarct ipsilateral and contralateral in the ALPS index between the poor and good prognosis groups. Abbreviations: DTI-ALPS, diffusion tensor image analysis along the perivascular space; AIS, acute ischemic stroke; ROI, regions of interest; FA, fractional anisotropy; NIHSS, National Institutes of Health Stroke Scale; NWU, net water uptake; mRS, modified Rankin Scale.

### 10. Impaired glymphatic clearance is associated with increased brain edema and poor functional outcomes in patients with acute ischemic stroke

We then investigated the relationship between impaired glymphatic clearance and brain edema and neurological outcomes. Results showed that the ALPS index of TIME 1 (**Fig. 9D**) and TIME 2 (**Supplementary Fig. 3A**) were linearly correlated with baseline NIHSS scores. However, there was no rank correlation between infarct volume measured from DWI images and ADC value and ALPS index (**Supplementary Fig. 3B-C**). Remarkably, brain edema detected by NWU of the ischemic lesion was linearly correlated with the ALPS index of TIME 2 (**Fig. 9E**).

During follow-up, 12 (66.7%) of 18 patients had 90-day good functional outcomes (mRS ≤ 2), and 6 (33.3%) had poor outcomes (mRS > 2). Patients with poor outcomes had significantly lower ALPS index (TIME 1) of both the infarcted and contralateral sides, compared with those with good outcomes (both *P* < 0.05; **Fig. 9F** and **Supplementary Table 5**). These results suggested that impaired glymphatic clearance measured by the decreased DTI-ALPS index was associated with increased brain edema and poor functional outcomes in patients with acute ischemic stroke.

## DISCUSSION

Here, we showed that the brain edema subsided on day 7 after MCAO when the BBB leakage kept increasing, indicating that the extravasation of protein-rich fluids into the brain was not temporally correlated with brain edema progression after MCAO. However, the time course of edema progression matched well the GS dysfunction after MCAO. When the GS function was enhanced by adrenergic receptor antagonists, brain edema was significantly alleviated on day 2 after MCAO, which was accompanied by reduced deposition of Aβ and improved cognitive function. Moreover, functional suppression of the GS delayed the absorption of brain edema, reflected by the recurrence of brain edema on day 7 after MCAO. In addition, patients with ischemic stroke represented significantly impaired glymphatic clearance from day 1 to day 3 after reperfusion, which was associated with increased brain edema and worse functional outcomes. These findings indicate that the GS is a key contributor to the formation of brain edema after ischemic stroke and targeting the GS may be a promising strategy for the treatment of post-stroke brain edema.

Generally, brain edema is divided into two distinct stages, the early stage of cytotoxic and ionic edema with an intact BBB, and the later stage of vasogenic edema with an impaired BBB. In the present study, we first examined the temporal evolution of brain edema after MCAO using MRI and electron microscopy and found that the brain edema developed as early as 6 h after MCAO, gradually peaked on day 2, then started to regress, and subsided on day 7. Our data are consistent with previous reports emphasizing the peak of vasogenic edema occurred 2-4 days after reperfusion(*12*). Intriguingly, we and other laboratories have previously demonstrated that the BBB is continuously open for the first week after reperfusion onset, and the leakage appears to further increase on day 7. These results raise a paradox that, at the late stage of ischemic stroke, brain edema starts to subside when the BBB is still leaky. In traditional opinion, this might be attributed to the rapid removal of edema fluid through the cerebrovascular endothelium(*4*). However, BBB opening after stroke is more conducive to the inflow of fluid to the brain parenchyma rather than the outflow of the fluid due to the hydrostatic and osmotic gradients(*28*).

Since the discovery of the GS as a brain-wide paravascular pathway providing a conduit for CSF influx and metabolic waste clearance, its role in the formation of brain edema has attracted widespread attention. A recent article described that within minutes of ischemic stroke, the rapid influx of CSF into the brain tissue drove acute tissue swelling, suggesting that ionic edema is caused by the influx of CSF, rather than by intravascular fluid as traditionally assumed(*18*). However, the specific role of the GS in the late stage of brain edema when the BBB is open has not been addressed. A prior study by Gaberel and colleagues(*29*) reported that the function of GS was impaired at 3 h post stroke, which was manifested as reduced CSF inflow into brain tissue. Subsequently, in a murine model of multiple microinfarcts, Wang et al.(*30*) reported that CSF tracer influx along the glymphatic pathway into the ipsilateral cortex was significantly reduced at 3 d post-microinfarct, with accompanying impairment of CSF tracer inflow into the contralateral hemisphere. Unilateral brain injury resulting in an impaired global influx of CSF along the glymphatic pathway has also been reported in traumatic brain injury(*25*), suggesting that the GS in both hemispheres of the brain is interdependent.

In the present study, we evaluated the effect of MCAO upon GS function by detecting the influx of CSF and the clearance of interstitial solute, with a focus on the edema peak on day 2 and edema regression on day 7. Consistently, by in vivo 2-photon observation, we found that CSF tracer influx into the ipsilateral cortex was almost abolished on day 2 after MCAO, while CSF tracer influx almost returned to normal on day 7 after MCAO. Similarly, by ex vivo fluorescence microscopy, we observed that CSF tracer penetration in the contralateral cortex was dramatically decreased on day 2 after MCAO and almost back to normal on day 7, although the CSF tracer dispersed broadly within the infarction area, possibly due to passive leakage from the disrupted BBB. These results demonstrated that CSF inflow along the glymphatic pathway was severely impaired on day 2 after MCAO, corresponding to the peak of brain edema, while CSF inflow was basically restored on day 7 after MCAO, corresponding to the regression of brain edema. More significantly, we found that patients with acute ischemic stroke showed impaired global GS clearance 1-3 days after reperfusion, prominently on the infarcted side, and gradually recovered thereafter, which was almost consistent with the clinical reports of edema peak in stroke patients(*31*). The impaired GS clearance was associated with increased brain edema and poor functional outcomes.

Dysfunction of the GS that impedes solute clearance has been reported in various neurological injuries, including subarachnoid hemorrhage, traumatic brain injury(*25*), status epilepticus(*32*), Alzheimer’s disease(*33*), and the aging brain(*34*). The GS may also play a positive effect in clearing edema fluids in days and weeks after stroke. In the present study, we found that the mice on day 2 after MCAO showed reduced glymphatic clearance of FITC-BSA and dextran tracer, as well as additional signs of glymphatic impairment such as low tracer signal of dCLNs and loss of AQP4 polarity. On the contrary, by observing the above indicators, the mice on day 7 after MCAO displayed normal function of glymphatic clearance. In addition, the whole-brain dynamic contrast-enhanced MRI imaging supported the notion that the glymphatic clearance of mice on day 7 after MCAO was significantly stronger than that on day 2 after MCAO. Combined with our MRI data on brain edema, the recovery of the GS function coincided with the recovery time of brain edema, suggesting that the GS pathway may be an important component of fluid outflow in the recovery stage of brain edema.

To further explore the effect of GS function on brain edema, we applied adrenergic antagonists and agonists to enhance or suppress the GS, respectively. As previously reported, the administration of adrenergic antagonists induced an increase in CSF tracer influx, and promoted increased lymphatic drainage of the CSF tracer, which was accompanied by improved AQP4 polarization distribution. Meanwhile, the brain edema on day 2 after MCAO was significantly alleviated, further supporting that the impaired function of GS plays an important role in leading to brain edema formation after MCAO. In contrast, adrenergic activation reduced the CSF tracer influx, and inhibited lymphatic drainage of the CSF tracer with recurrent brain edema at day 7 after MCAO, suggesting that the GS plays an important role in mediating the clearance of brain edema.

The mechanisms behind brain-regulated suppression of glymphatic activity by adrenergic inhibition may include the following aspects. Firstly, several studies have verified that inhibition of adrenergic signaling increased the interstitial volume, which is closely related to the function of GS. Regrettably, the changes in interstitial volume were not examined in the present study.

Secondly, the surface expression of AQP4 must be polarized to astrocytic-endfeet abutting cerebral vessels for efficient glymphatic clearance. As we observed, reduced AQP4 polarity after MCAO was partially reversed by adrenergic inhibition (**Supplementary Fig. 4**). We speculate that promoting the recovery of AQP4 polarization may also be one of the factors. However, the exact regulatory mechanisms need to be further studied.

In conclusion, our data demonstrated for the first time, that the progression and resolution of brain edema after MCAO coincided with the impairment and recovery of GS function, and intervention of the GS function could significantly modulate the extent of brain edema, suggesting that GS transport may play a decisive role in the progression of brain edema after ischemic stroke, even in the presence of BBB leakage. Further investigations on the mechanism of regulating the GS function are needed to seek clinically effective molecular targets for the prevention and treatment of brain edema

## MATERIALS AND METHODS

### Study approval

All animal experiments in this study were approved by the Animal Care and Use Committee of the Nanfang Hospital, Southern Medical University (Guangzhou, China), and adhered to the National Institute of Health Guide for the care and use of laboratory animals (NO. NFYY-2018-0937). The study of stroke patients was performed following the Declaration of Helsinki and was approved by the Medical Ethics Committee of Nanfang Hospital (NO. NFEC-2019-189). Written informed consent was obtained in all subjects.

### Animals

Male C57BL/6J background mice were obtained from the Experimental Animal Center of Southern Medical University. All mice were housed under a 12-h light/dark cycle with free access to water and standard chow. All efforts were made to minimize the number of animals used and the pain inflicted on them.

### MCAO model and drugs

The 8 to 10 weeks old mice were subjected to 45-min transient middle cerebral artery occlusion (MCAO), followed by up to 7 days of reperfusion, as we described previously(*10*). Successful modeling of MCAO was defined as a reduction of focal cerebral blood flow by more than 75% of baseline and full recovery of blood flow after reperfusion, which were monitored by a laser Doppler flowmetry (Moor Instruments, Wilmington, United States). At 30 min after reperfusion, the MCAO mice were randomly allocated to different groups. For adrenergic inhibition, a cocktail of adrenergic receptor antagonists (prazosin, 1mg/kg, atipamezole,1 mg/kg and propranolol,10 mg/kg) was intraperitoneally used once daily until sacrifice. For adrenergic stimulation, ephedrine (30 mg/kg, Macklin) was subcutaneously injected once daily from day 3 to day 7 after MCAO induction. Mice in the control group received an equivalent volume of saline (**Supplementary Fig. 5**).

### Magnetic resonance imaging (MRI) experiments in mice

Brain MRI on mice was performed as described previously(*10*). After 4% isoflurane induction anesthesia, the mice were transferred to the MRI-compatible cradle and maintained anesthesia with 1.5% isoflurane. The brains were scanned by a 7.0T Pharmascan MRI system (Bruker, Germany), and fifteen contiguous coronal images of each mouse were collected. Edema formation was visualized from T2-weighted images and diffusion-weighted images (DWI) and semi-quantified as midline-shift (MLS) and relative ADC value (rADC) on restructured ADC images, respectively. MLS was calculated using the following formula: MLS = (A−B)/2, where A and B represent the distance between the cortex outer border of the ipsilateral side and contralateral side and the middle of the third ventricle, respectively. rADC% was expressed as (ADC_ips_/ADC_con_) × 100%, where the corresponding ADC value was measured in the ipsilateral ischemic and contralateral symmetric regions.

Contrast-enhanced MRI was used to visualize the brain-wide GS function(*20*). Gd-DTPA (6 μL) was microinjected into the cisterna magna of mice at a rate of 1.6 μL/min. This infusion rate was chosen because it did not affect intracranial pressure(*35*). High-resolution T1-weighted imaging was performed before Gd-DTPA injection (baseline) and thereafter at 0.5, 2, 4, and 6 h after Gd-DTPA injection. The above data were calculated by a researcher blinded to group allocation using Image Display and Processing software (Bruker).

### BBB permeability assay

Evans blue (EB) dye was performed as described previously(*10*). EB (2% in saline, 4 mL/kg, Sigma-Aldrich) was injected via the tail vein and allowed to circulate for 1 h. Then, the mice were transcardially perfused with saline and the brains were removed to detect the contents of EB.

### Intracisternal CSF tracer infusion

Intracisternal injection was performed as previously described(*32*). Briefly, anesthetized mice were fixed in a stereotaxic frame and the cisterna magna was surgically exposed. Using a syringe pump (RWD Life Science, China) with a Hamilton syringe connected to a 30-gauge needle, 6 μL of EB (1% in artificial cerebrospinal fluid) was infused at a rate of 1.6 μL/min. After injection for 30 min, the animals were sacrificed and the brain was removed and fixed overnight by immersion in 4% paraformaldehyde (PFA) in PBS. Brain sections were cut and imaged on an Olympus FV10i fluorescence microscopy. Tracer influx was quantified by a blinded investigator using ImageJ software.

### Parenchymal clearance assay

Mice were anaesthetized with 2% isoflurane and and positioned in a stereotaxic frame. For intrastriatal injection, the skin was opened and the skull was exposed. A burr hole was made with a hand-held drill on the coordinates: +0.5 mm anterior/posterior, +2.0 mm medial/lateral, and +3.25 mm dorsal/ventral from bregma. One μL of 1% fluorescein isothiocyanate-conjugated bovine serum albumin (FITC-BSA, MW 66 kDa) was injected at a rate of 0.1 μL/min via the micropump. The micropipette remained in place for 5 min and then was withdrawn very slowly to avoid any possible backflow. Two hours after intrastriatal injection, mice were transcardially perfused with a fixative containing 4% PFA in 0.1 M PBS. The brains and dCLNs were removed and post-fixed for 24 h.

### In Vivo 2-Photon Fluorescence Imaging

CSF tracer penetration into the brain parenchyma was visualized through an artificially thinned cranial window using a 2-photon laser scanning (Olympus) microscope, as described previously(*32*). Animals were anesthetized, and 6 μL of fluorescein isothiocyanate-dextran (FITC-dextran; 70 kDa, 1% dissolved in artificial CSF; Sigma-Aldrich) was microinjected into the cisterna magna to evaluate the GS influx function 30 min before imaging. To detect the glymphatic clearance after MCAO, intracortical injection of 1 μL of 1% 40 kDa Rhodamine B isothiocyanate-dextran (RITC-dextran, Sigma-Aldrich, dissolved in artificial CSF) was performed. Corresponding tracers (0.2 mL, 1% dissolved in saline; Sigma-Aldrich) were injected intravenously to visualize the vasculature immediately before imaging. An Olympus 2-photon imaging system (Fluoview FV1200) equipped with a water immersion objective (25 ×, 1.05 NA) was used for imaging.

### AQP4 immunohistochemistry and quantification

AQP4 polarization was evaluated as previously reported(*34*). Higher perivascular AQP4 immunoreactivity and lower parenchymal AQP4 immunoreactivity indicated increased polarization and vice versa. First, we measured the median immunofluorescence intensity in the perivascular area. Then, the percentage of the region exhibiting AQP4 immunofluorescence greater than or equal to perivascular AQP4 immunofluorescence (AQP4% Area) was measured by threshold analysis. AQP4 polarization was expressed as the percentage of the region that exhibited lower AQP4 immunoreactivity than the perivascular end-feet (polarization=100-AQP4% Area).

### Ligation of deep cervical lymph nodes (dCLNs)

On day 3 after MCAO, the mice underwent dCLNs ligation. In brief, after anesthesia, a longitudinal sterile incision was made on the neck from the mandible to the sternum. The muscles and fascia were carefully separated along the trachea using blunt-tipped forceps under the stereomicroscope to expose the dCLNs, which were located below the sternothyroid muscle. The afferent and efferent vessels were then carefully ligated using 6-0 nylon sutures. Sham-operated mice were only surgically exposed dCLNs without ligation.

### Morris Water Maze Test

The Morris water maze was carried out to evaluate short-term spatial learning and memory. Briefly, the mice were first examined for spatial learning during 5 consecutive days of hidden platform training with four trials per day. Then, the platform was removed and each mouse was tested for spatial memory on a single 60-s probe trial. Swim paths were recorded by the TSE VideoMot2 video tracking system (TSE Systems GmbH, Bad Homburg, Germany). The number of crossings over the former platform location and the time spent in the target quadrant during the probe trial were analyzed.

### Measurement of diffusion tensor image analysis along the paravascular space (DTI-ALPS) in patients

We retrospectively enrolled 18 adult patients with acute ischemic stroke who had diffusion tensor imaging (DTI) examination performed around 24 hours after endovascular treatment (EVT) from a prospective cohort of biomarker studies in patients undergoing EVT for acute ischemic stroke in Nanfang Hospital, Southern Medical University (ChiCTR No. ChiCTR2200057845).

All MRI data including DTI and DWI were acquired on a 3.0 T scanner (uMR 780, United Imaging Healthcare, Shanghai, China) with a 32-channel head coil for signal reception. DTI-ALPS was used to evaluate the glymphatic clearance function(*27*). In brief, the main difference between the x-axis diffusivity in both projection and association fibers (i.e., Dxxproj and Dxxassoc) and the diffusivity that was perpendicular to the x-axis but not parallel with major fibers (i.e., Dyyproj and Dzzassoc) from DTI images were regarded as the existence of the PVS. The glymphatic clearance activity was quantified using the DTI-ALPS index defined by Taoka et al. (*27*) as follows:

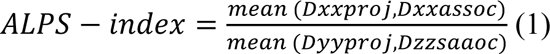

For all participants, the DTI-ALPS indexes on the infarcted side and the contralateral side were calculated respectively.

### Human clinical and imaging information

Human clinical information was extracted from the electronic database, which included age, sex, the National Institutes of Health Stroke Scale (NIHSS) score at admission, a subtype of stroke, occlusion vessel, infarct hemisphere (left or right), time from onset to puncture/recanalization, and 90-day modified Rankin Scale (mRS). The 90-day mRS of 0-2 and 3-6 were defined as good and poor prognoses, respectively. Brain edema was quantified by net water uptake (NWU) of the ischemic lesion, which was calculated from follow-up NCCT at 72 hours after EVT using the density values of the hypodense infarct area (D_infarct_) and the corresponding area on the contralateral side (D_normal_) according to the following formula(*36, 37*):

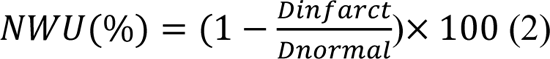

### Statistics

For mice experiments: all data are expressed as the mean ± standard deviation (SD). Statistical differences among multiple groups were compared using one-way analysis of variance (ANOVA) followed by Tukey’s multiple comparisons test. The differences between the two groups were analyzed using unpaired t-tests. The MRI data for monitoring brain edema changes were examined by one-way repeated measurement ANOVA followed by Dunnett’s multiple comparisons test. The two-photon imaging and contrast-enhanced MRI data of glymphatic clearance at different time points were analyzed by two-way repeated measurement ANOVA, and multiple comparisons were made by Sidak’s test and Dunnett’s test respectively. Statistical analysis was performed using GraphPad Prism version 9.0 (GraphPad, La Jolla, USA). Statistical significance was set at *P* < 0.05.

For clinical data: the Shapiro-Wilke test was used to determine if continuous variables were normally distributed. Normal distribution variables were expressed as mean ± standard deviation (SD), while non-normal distribution variables were expressed as median (interquartile range, IQR). The inter-observer reliability of the ALPS index was determined by intraclass correlation coefficients (ICCs) with a 95% confidence interval (CI). The final values of the ALPS index were calculated by the mean value of two observers.

Interhemispheric differences in ALPS index values between the infarcted side and the contralateral side at the same time point were compared using a paired-samples *t*-test. The overall analysis of the ALPS index of the ipsilateral and contralateral side of the infarct that was measured repeatedly at different time points using two-way repeated measurement ANOVA, then combined with paired samples t-test and Least Significant Difference (LSD) methods for two-by-two comparisons at different times or on both sides of the infarct. Independent samples t-test was used to assess the difference in ALPS index values between the good and poor prognosis groups. Bivariate correlations between baseline NIHSS, infarct ADC, infarct volume, infarct NWU, and ALPS index of both the infarcted side and the contralateral side were determined using Pearson’s correlation analysis or Spearman correlation analysis as appropriate. SPSS 25.0 (IBM, Armonk, United States) and GraphPad Prism were used for analyzing data and graphing pictures. *P*-values < 0.05 (two-tailed) were considered to be statistically significant.

## Data Availability

All data are available in the main text or the supplementary materials.

## List of Supplementary Materials

Material and Methods

Results

Table S1 to S5: Data from clinical patients

Fig S1 to S6: Data from clinical patients and animal study

References

## Acknowledgments

We are extremely grateful to patients and their families, the Department of Biostatistics of Southern Medical University for technical support.

## Author contributions

JZ, K. Liu, Z. Li, Y. He, YC, C. Lin, M. Yu performed animal experiments and analysis. J. MO, Q. Chen, Y. Xu, X. Tan performed human experiments and analyzed results. JZ prepared the manuscript with input from all authors. K. Huang and S. Pan conceived and designed the studies.

## Data and materials availability

All data are available in the main text or the supplementary materials.

